# Public Health response to an outbreak of SARS-CoV2 infection in a Barcelona prison

**DOI:** 10.1101/2020.11.08.20227744

**Authors:** A Marco, C Gallego, V Pérez-Cáceres, RA Guerrero, M Sánchez-Roig, RM Sala-Farré, J Fernández-Náger, E Turu

## Abstract

An outbreak of SARS-CoV2 infection in a Barcelona prison was studied after seven cases appeared in nine days. One hundred and eighty-four people (148 inmates and 36 prison staff) were evaluated by rt-PCR. Thirty-nine (24.1%) were positive: 33 inmates and six staff members. The inmates were isolated in prison module 4, which was converted into an emergency COVID unit. Two people (one inmate and one health worker) were admitted to hospital for clinical deterioration. There were no deaths. Outbreaks pose a huge risk, must be detected early, are difficult to manage, and require optimal coordination between health and prison authorities.

## 1. Background

On 14 March, a state of emergency was declared in Spain due to the spread of the SARS-CoV-2 infection. In prisons, mobility and interpersonal contacts were severely restricted, with the suspension or reduction of prison activities, communications, and permits. A 14-day confinement was also ordered for new admissions. These restrictions were approved by a Ministerial Order. The government of Catalonia (an autonomous community in Spain) has responsibility for health and prison policy throughout the region. In Catalonia, 8,300 inmates are held in nine prisons and in five open penitentiary centers. Prison medical services, health programs and healthcare circuits depend on the public health system managed by the Catalan Institute of Health. Up until mid-April, in order to reduce the population exposed, the Catalan government released 17% of the prison population (n = 1,425 inmates). This figure was considerably higher than the average of the countries of the European Union, which was 5.1% [1]. Seventeen days after adopting these measures, when 13 cases had been diagnosed in four other prisons the first cases of SARS-CoV2 Infection were detectd in Quatre Camins Prison (henceforth, QCP).

## 2. Outbreak detection

Quatre Camins Prison is located in La Roca del Vallés, in the province of Barcelona. It houses 946 inmates, all male, in 14 residence modules plus an extra module for new entrants, and a nursing department.

Between 31 March and 9 April, QCP reported seven cases in inmates in module 4 (MR4) of the prison. Table 1 displays the descriptive characteristics of these seven cases and their time spent in prison. For this reason, the decision was taken to study all the inmates admitted to QCP MR4, as well as the health workers and non-health workers who had had close contact with this population group. Family members were not studied because there had been no contact with people from outside the prison since 15 March.

**Table 1.** Descriptive characteristics and incarceration time of the seven initial cases detected in QCP MR4 between 31 March and 8 April, 2020.

| Case | Country of birth | Age | HIV | Time in prison (days)<sup>a</sup> |
| --- | --- | --- | --- | --- |
| 1 | Philippines | 63 | No | 761 |
| 2 | Dominican Republic | 35 | No | 65 |
| 3 | Ecuador | 27 | No | 591 |
| 4 | Spain | 32 | No | 437 |
| 5 | Spain | 31 | No | 2,999 |
| 6 | Spain | 30 | No | 203 |
| 7 | Spain | 32 | No | 1,401 |

QCP: Quatre Camins Prison; MR4: Residential Module 4.
In QCP MR4 and at the time of diagnosis (rt-PCR +)<sup>a</sup>

For the study of the outbreak, “close contacts” were identified, using the definition of the Spanish Ministry of Health: that is, in the prison context, people who had had contact with the case in the 48 hours before the onset of symptoms (or diagnosis, in the case of asymptomatics) until the moment when the case was isolated.

## 3. Population screened

On 9 April 2020 the MR4 inmates and the staff who had been in close contact with them in the past 14 days were administered the real-time reverse transcription polymerase chain reaction test (rt-PCR) with samples of nasopharyngeal/oropharyngeal exudate. All the samples were analyzed at the laboratory of the Germans Trias University Hospital and were categorized into two groups: a) symptomatic cases, or b) close non-symptomatic contacts.

Although the care of health workers is also managed by the Catalan Institute of Health, it is overseen by a different department and access to health workers’ data is restricted. For this reason, the rt-PCR was performed in health workers who might have had contact with MR4 inmates, but if the infection was confirmed, the worker was referred for control and follow-up by his/her own healthcare network. Pending the results of the rt-PCR, the inmates were left in isolation in MR4.

184 subjects were screened: a) 148 inmates (145 inmates in MR4 and three more from that module but who were in isolation in the Nursing Department due to their previous close contact with inmates diagnosed with the infection); b) 31 non-health workers, and c) five health workers: one doctor, one nurse and three health assistants.

One hundred forty-six of the inmates screened were asymptomatic and two had mild symptoms (one case with ageusia and anosmia and another with low fever and general discomfort). Only one of the health workers presented clinical symptoms (fever, cough, and moderate respiratory distress). All non-health workers were asymptomatic.

## 4. Data of the population screened

In addition to the seven cases already diagnosed, 39 more were positive on rt-PCR screening: three symptomatic and 36 asymptomatic subjects. The positive rt-PCR rate was therefore 24.1%. Figure 1 shows the distribution according to the group studied, the rt-PCR results, and the presence/absence of symptoms.

**Figure 1.**
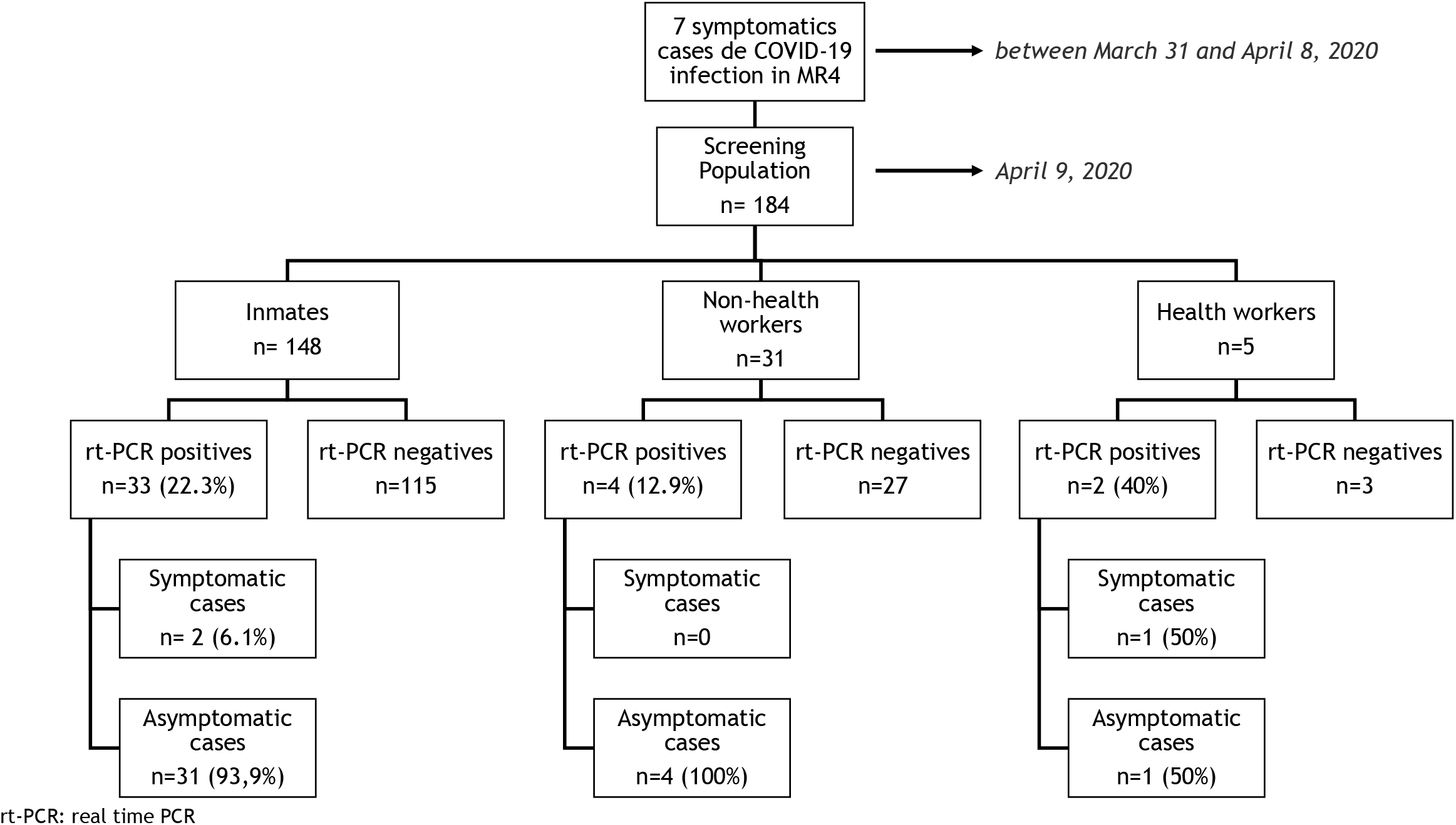
Distribution according to population group, rt-PCR result and symptoms or not.

Regarding clinical evolution, only two individuals (one inmate and one health worker), were admitted to the hospital, but neither required intensive care. All 39 patients evolved satisfactorily.

All inmates were men with a mean age of 40 ± 7.3 years (range 21-76 years). Seven (4.7%) were ≥ 60 years old and 52 (35.1%) were not Spanish. Regarding chronic diseases, eight (5.4%) had diabetes (one with associated heart disease) and 13 (8.8%) were infected with HIV. Immune status, the combination of antiretroviral treatment and other characteristics of the HIV-infected inmates screened are presented in Table 2.

**Table 2.**
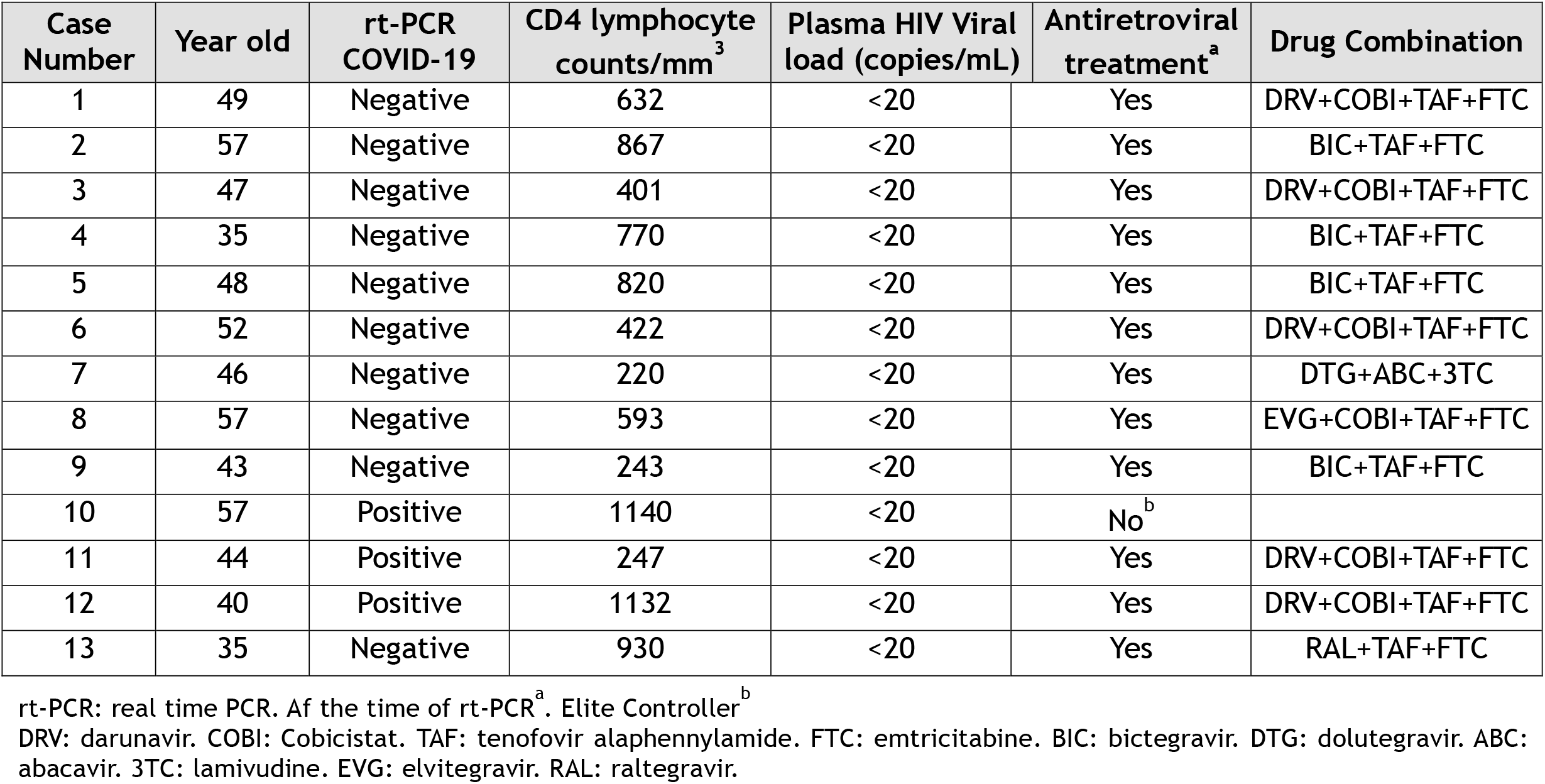
Characteristics of HIV-infected individuals undergoing screening.

There were no statistically significant differences between inmates with positive or negative rt-PCR in terms of age (mean 39.2 vs. 40.6 years; p = 0.89), history of diabetes (25% vs. 22.1%; p = 0.40), or HIV infection (9.1% vs. 8.7%; p = 0.91). According to origin, however, significant differences were found, since more people of Latin American, origin had positive rt-PCR (78.6% vs. 16.4% in those of other origins; p <0.001; OR = 18.67, 95% CI: 4.81-72.43). Figure 2 shows the proportion of positive rt-PCR according to the origin of the patients.

**Figure 2.**
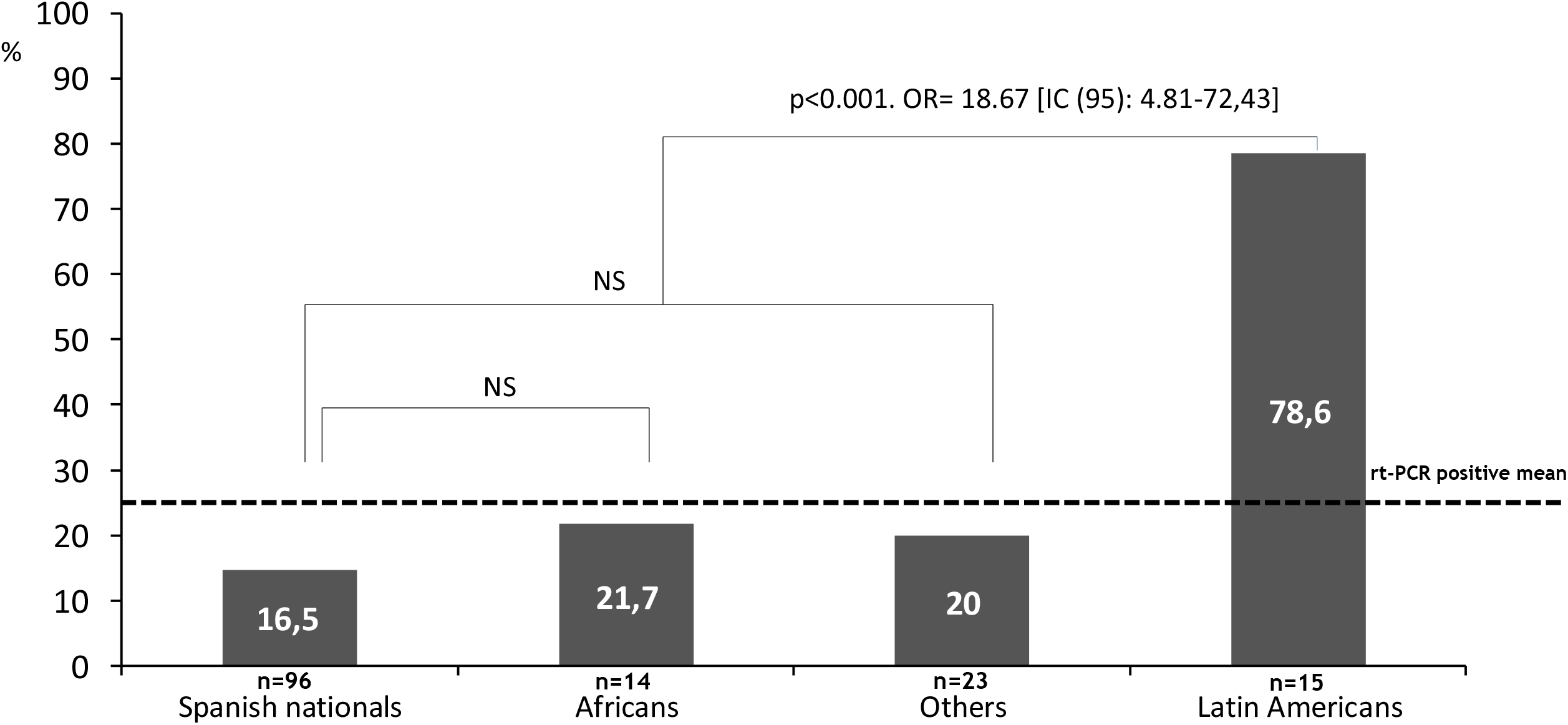
Distribution of positive rt-PCR according to the origin of the studied patients.

## 5. Outbreak control measures

Isolation at home was recommended in the six workers with positive rt-PCR tests, and the result was reported to the Occupational Risk Prevention Unit. The subsequent control and follow-up were carried out by their corresponding medical services. As for inmates, those with positive rt-PCR were isolated in MR4, which was disinfected sequentially by zones. Inmates with negative rt-PCR were confined in MR1.

MR4 was considered an emergency COVID-19 unit, created because of the number of infected inmates who were asymptomatic or mildly symptomatic but did not present criteria for hospitalization. The unit adopted a series of organizational and functional measures to guarantee the safety, quality and efficiency of the care given to these low complexity cases. Cleaning, laundry, waste management and the distribution of food and medication were organized according to the recommendations of the Catalan Health Service [2].

The following controls were imposed: a) strict isolation of the unit, which only key health and non-health workers were authorized to enter and leave; b) use of individual protection equipment; and c) clinical controls (oxygen saturation, temperature, and enquiries about the appearance of symptoms) twice daily. During the stay in isolation, only one of the 33 inmates admitted was hospitalized (due to clinical deterioration).

The inmates with negative rt-PCR were transferred to MR1, where they were placed in confinement. There they were allowed to share some spaces in small groups, but wearing a mask at all times. The aim was to ensure that they were not incubating the SARS-CoV2 infection or were rt-PCR “false negatives”. Therefore, in this a situation of confinement clinical controls (oxygen saturation, temperature, and enquiries about the appearance of suspicious symptoms) were performed twice daily. No inmate presented fever or any clinical suspicion of COVID-19 during the period of confinement.

Just over half (51.5%) presented a negative rt-PCR after 14 days of isolation, and 81.8% at 21 days, while 18.2% were negative only after four weeks.

## 6. Discussion

This report of the outbreak at the QCP is one of the first descriptions (if not the first) of COVID-19 in the prison setting in Europe. SARS-CoV2 infection was detected in 40 inmates and six workers (24.1%) of the individuals studied. This rate is high, though below the 30% observed in a long-term care nursing center [3], and below the 35% recorded in an outbreak in a hospital [4] (although the latter report corresponded to the first indigenous case of COVID-19 infection in the US, so it was unsuspected and exposure was increased by the use of multiple aerosol generation procedures).

The rate reported in this outbreak is higher than that observed in the Diamond Princess Cruise passengers after 14 days of quarantine in February 2020, which was 20.7% [5]. It has been calculated that in the cruise ship the R0 (the mean number of people who will contract a disease from one contagious person) was 5-14 times higher than the normal figure of 1.5-3.0, because of the high occupant density and confined space [6]. A similar process, even more intense, may have occurred in the QCP MR4. SARS-CoV2 spreads widely in closed spaces and this is probably the reason why 24.1% of the admissions in MR4, and 78.6% of the Latin American inmates, became infected. In prisons, members of racial and ethnic minorities tend to stick together and protect each other, and share cells, activities, and even food. There is no greater genetic predisposition to infection in ethnic or racial groups. Therefore, the expansion of the infection and the very high positive rt-PCR rate in the Latin American inmates were presumably due to the close contact they maintain with each other.

It was not possible to identify the index case. In this infection, the mean incubation time is 5.1 days, but 97.5% of symptomatic cases occur within 11.5 days of exposure [7], so the index case may have been asymptomatic or one of the initial seven cases. The lack of cases prior to the outbreak and restrictions regarding contact with the outside world (no prison leave, no visits, and 14-day confinement of new admissions) suggest that the transmission may have started from an asymptomatic case, either a prisoner or (more likely) a member of the prison or health staff. In fact, it has been estimated that silent disease transmission during the presymptomatic and asymptomatic stages is responsible for more than 50% of the overall attack rate in COVID-19 outbreaks [8].

Regarding age, the subjects affected were young (mean age 40 years) and many had no relevant medical history. As is customary in these patients, the evolution was satisfactory and no deaths were recorded. The case fatality rate of zero is a very important finding and needs to be emphasized. In addition to the younger age of the patients, the highly effective measures adopted probably had a bearing on the results.

It should be remembered, however, that 9.1% of the inmates infected in the outbreak had HIV infection, even though all were virologically controlled and all had a CD4 lymphocyte count/mm^3^ above 200. Currently, there are no solid data to demonstrate that HIV-infected individuals with COVID-19, present poorer clinical evolution if they are well controlled and have moderately preserved immunity [9].

It should also be noted that 94.8% of subjects with positive rt-PCR did not present symptoms. One review estimated that between 40% and 45% of the cases of SARS-CoV-2 infection are asymptomatic [10]. As recently suggested, asymptomatic transmission is probably the Achilles heel of the pandemic [11]. Asymptomatic patients transmit the infection silently and may interact more with other people because they do not feel sick [8, 12]. In accordance with reports of other outbreaks [13,14], our results confirm that the isolation of both symptomatic and non-symptomatic patients and the study of all contacts is essential in order to control the outbreak.

In short, the outbreak presented here shows that outbreaks of SARS-CoV2 infection in closed environments are a real possibility. They pose a huge risk, must be detected early, are difficult to manage, and require optimal coordination between the health and prison authorities. When they occur, general screening by means of PCR and the isolation and evaluation of those infected are key measures. Symptom-based surveillance must be supplemented by rapid contact-based monitoring in order to avoid asymptomatic spread among prisoners, health workers, and ultimately in the community at large.

## Data Availability

I declare the availability of all the data referred to in the manuscript

## Acknowledgments

We thank all the members of the health team at the QCP. Without their efforts, the outbreak could not have been brought under control.

